# Unveiling the Epigenetic Impact of Vegan vs. Omnivorous Diets on Aging: Insights from the Twins Nutrition Study (TwiNS)

**DOI:** 10.1101/2023.12.26.23300543

**Authors:** Varun B. Dwaraka, Lucia Aronica, Natalia Carreras-Gallo, Jennifer L Robinson, Tayler Hennings, Aaron Lin, Logan Turner, Ryan Smith, Tavis L. Mendez, Hannah Went, Emily R. Ebel, Matthew M. Carter, Erica D. Sonnenburg, Justin L. Sonnenburg, Christopher D. Gardner

**Affiliations:** TruDiagnostic, Inc. 881 Corporate Dr. Lexington, KY, 40503; Stanford Prevention Research Center, Department of Medicine, School of Medicine, Stanford University, Stanford, CA 94305, USA; Seattle Children’s Research Institute, Seattle, WA, 98101, USA; Department of Microbiology and Immunology, School of Medicine, Stanford University, Stanford University, Palo Alto, California; Chan Zuckerberg Biohub, San Francisco, California; Center for Human Microbiome Studies, Stanford University School of Medicine, Stanford, California

**Keywords:** Diet and Nutrition, Epigenetic clocks, Aging, Epigenome Wide Association Study, Vegan, Omnivore

## Abstract

Geroscience has emerged as a field focusing on interventions to attenuate molecular changes associated with aging. While lifestyle modifications, medications, and social factors are recognized influencers of the aging process, a comprehensive understanding of the intricate molecular mechanisms necessitates an in-depth exploration of the epigenetic landscape. Notably, the specific epigenetic clock and predictor effects of a vegan diet, compared to an omnivorous counterpart, remain inadequately explored, despite indications of potential impacts on aging-related outcomes. This study addresses this knowledge gap by examining the impact of an eight-week entirely plant-based or healthy omnivorous diet on blood DNA methylation in paired twins. Results show distinct responses, with the vegan cohort solely exhibiting significant decreases in overall epigenetic age acceleration (PC GrimAge, PC PhenoAge, DunedinPACE), including among specific systems (Inflammation, Heart, Hormone, Liver, and Metabolic), aligning with anti-aging effects of plant-based diets. Analyses of methylation surrogates of clinical, metabolite, and protein markers indicate diet-specific shifts, while exemplifying DNA methylation markers in predicting complex traits influenced by diet. Comprehensive epigenome-wide analysis unveils diet-specific differentially methylated loci, offering insights into influenced pathways. This study sheds light on the advantageous aging benefits of a healthy vegan diet, while providing a foundation for future personalized interventions using epigenetic age clocks in promoting overall well-being.

## Introduction

While advances in technology and medicine have allowed the average person to live longer, age-related disease and impairment remain an issue that greatly impacts individuals, healthcare systems, and consequently, society. Aging is associated with increases in health care costs and financial stress for social insurance systems (Jin et al. 2014). In light of these challenges, the field of geroscience has emerged, proposing interventions aimed at slowing down or reversing the molecular changes that occur with aging. These interventions encompass a wide range of factors, including lifestyle modifications, nutrition, medications, sleep, and social factors, all of which can influence the aging process and potentially delay or prevent the onset of multiple chronic diseases, ultimately extending a healthy lifespan (Bauman et al. 2016; Kennedy et al. 2014; Campisi et al. 2019). Consequently, the exploration of nutritional and dietary recommendations has become an increasingly significant area of research within the broader field of aging, providing insights into how dietary choices can impact the aging process and overall health outcomes.

However, unraveling the intricate molecular mechanisms through which diets influence aging necessitates a deeper understanding of the epigenetic landscape (Sen et al. 2016). Epigenetic modifications, such as DNA methylation, have emerged as pivotal regulators of gene expression and provide a promising avenue for investigating the effects of vegan diets on the aging processes (Miles et al. 2020). The epigenetic effects of a vegan diet, in comparison to an omnivore diet, remain largely unexplored, with limited available evidence. Although certain studies have indicated potential positive impacts of specific components of a vegan diet, such as heightened intake of vegetables and fruits, on epigenetic aging, concerns have been raised regarding potential deficiencies in essential “epi-nutrients” necessary for effective epigenetic regulation (Craig 2010). Notably, nutrients including vitamin B12, vitamin D, choline, omega-3 fatty acids, and zinc are among the concerns associated with a vegan diet, as their availability may be compromised. In addition to these concerns, other work on diets has aimed to discover the association between diets and longevity (Trichopoulou and Vasilopoulou 2000; Roberts et al. 2017). Moreover, other research on diets has shown the effect of a Mediterranean diet on frailty (Capurso et al. 2019). This also corresponds to other studies exploring protein intake and age on the onset of frailty (Coelho-Junior et al. 2020).

Epigenetic clocks, derived from DNA methylation patterns, have emerged as powerful tools for estimating biological age and predicting age-related outcomes. These clocks have also been refined over time to incorporate known clinical factors, making them sensitive and reliable indicators of aging-related changes (Higgins-Chen et al. 2022). Additionally, epigenetic interpretation algorithms have proven valuable in predicting relative immune cell levels and protein expressions, providing insights into immune system functionality through immune deconvolution (McCartney et al. 2018; Chen et al. 2023; Luo et al. 2023). Moreover, these clocks can estimate the number of cell cycle divisions, reflecting cellular senescence and potential disease susceptibility (Teschendorff 2020).

While aging intervention studies have the added challenge of requiring long enough periods of time for the interventions to show a statistically significant effect, advancements in DNAm-based analysis, such as phenotypically and clinically trained DNAm clocks, have allowed for changes in the pace of aging and risk factors related to aging to be studied (Rutledge, Oh, and Wyss-Coray 2022). Epigenetic age trials using these epigenetic clocks have discovered that different diets such as a Mediterranean diet and DASH diet have shown improvement in certain aspects related to aging (Gensous et al. 2020; Galkin et al. 2023). In particular, a Mediterranean diet has been shown to both slow aging and delay the onset of frailty (K. Kim et al. 2022).

Given the discussion on which diets are most beneficial to longevity, this study aims to discover the effect of an eight-week entirely plant-based or healthy omnivorous diet on DNAm of blood in paired twins and evaluate risk factors related to age as well as other biomarkers related to health.

## Results

### Participant Attrition and adherence to protocol

To investigate the impact of diet on the methylome, blood samples from a randomized clinical trial were used to quantify methylation (Landry et al. 2023). As shown in **Figure 1**, to quantify methylation, whole blood was collected to establish a baseline measure of methylation at the time of starting the trial (Week 0) and at the conclusion of the study (Week 8). Baseline characteristics by diet group appear in **Table 1**. Among 21 pairs of twins, the randomized mean age was 39.9 (SD 13) years, 77.3% were women, and the mean body mass index was 26 (SD 5). The BMI of both cohorts was largely equivalent (average Vegan BMI = 26.3, average Omnivore BMI = 26.2).

**Figure 1.**
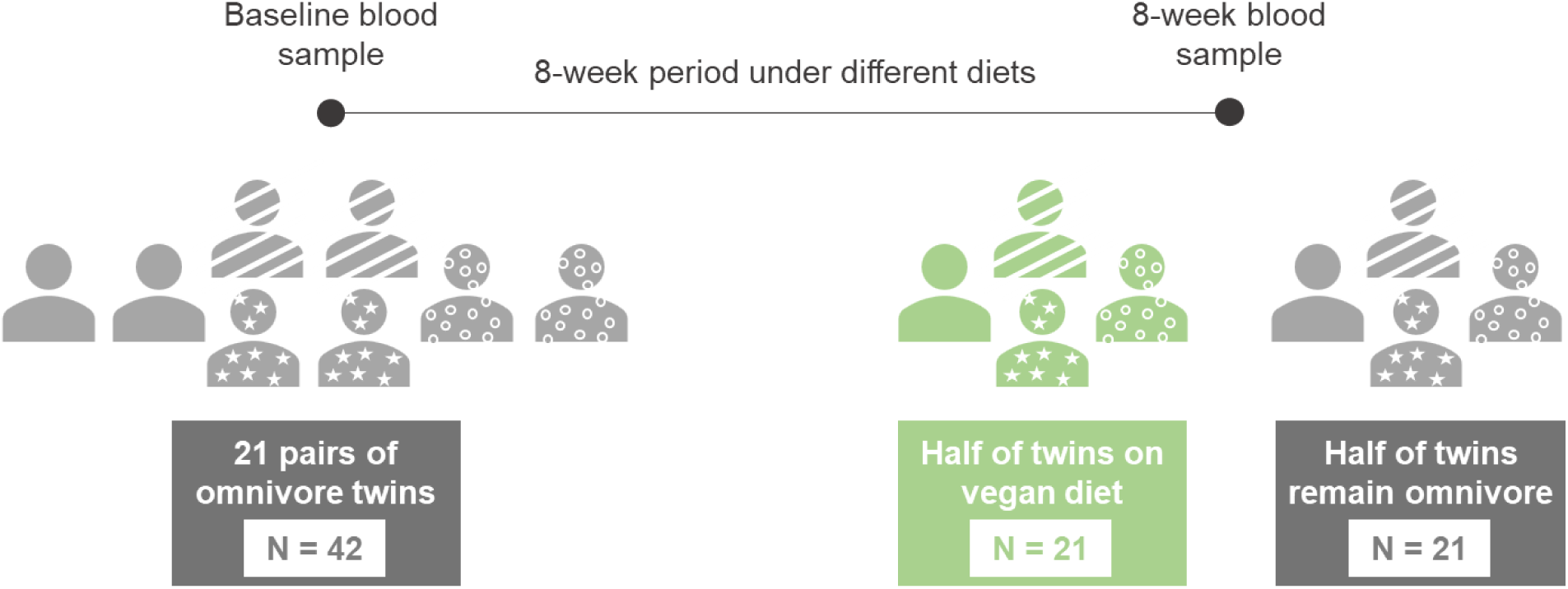
**Timeline diagram for the study design**. A total of 21 pairs of twins (N=42) were subjected to a vegan diet (N = 21, labeled in green) and an omnivore diet (N = 21, labeled in orange). Blood was collected for Baseline at the start of the trial (Week 0) and at the end of the trial (Week 8) and methylation states were quantified using the EPIC 850k array.

**Table 1.** Baseline Characteristics of the Study Participants.

|  | Vegans | Omnivores |
| --- | --- | --- |
| **Sample Size (N)** | 21 | 21 |
| **Males (%)** | 5 (18.5%) | 5 (18.5%) |
| **Females (%)** | 16 (76.2%) | 16 (76.2%) |
| **Mean Age (SD)** | 39.9 (13) | 39.9 (13) |
| **Mean BMI (SD)** | 26.3 (5) | 26.2 (5) |

### Diet type impacts changes in epigenetic age and telomere length

To investigate the response to diet upon biological age and telomere length, we quantified and analyzed several biological age and telomere length predictors derived from DNAm. These included the principal component (PC) based clocks: the first generation multi-tissue Horvath (Horvath1) and skin+blood Horvath (Horvath2); and the second generation PhenoAge, GrimAge, and telomere length (DNAmTL) clocks. Additionally, several non-PC clocks were included as well: the first generation Zhang clock based on the elastic net (Zhang-EN) and BLUP (Zhang-BLUP) method; the second generation multi-omic informed OMICmAge, and the third generation DunedinPACE clock. To better understand the impact of diet on the epigenetic age of specific organ systems, we also calculated the individual ages of 11 organ systems: Heart, Lung, Kidney, Liver, Brain, Immune, Inflammatory, Blood, Musculoskeletal, Hormone, and Metabolic. In addition, a composite age of the system was also calculated as Systems Age. In the vegan group, we observed significant decreases in the following epigenetic age metrics: PC GrimAge (mean delta EAA = -0.3011, p = 0.033), PC PhenoAge (mean delta EAA = -0.7824, p = 0.014), and DunedinPACE (mean delta PACE = -0.0312, p = 0.00061) significantly decreased at 8 weeks relative to 0 weeks (**Figure 2A-C**). Similarly, we observed significant reductions in the composite systems age metric age, which was corroborated by significant reductions of 5 out of 11 systems: Inflammation, Heart, Hormone, Liver, and Metabolic (**Figure 2D-2I**). In contrast, no epigenetic clock or telomere measure exhibited significant changes in the omnivorous cohort, suggesting that the omnivorous diet did not induce any epigenetic age methylation changes. Taken together, these findings suggest that the observed DNA methylation changes may contribute to overall decreases in epigenetic age in response to a vegan diet, which is not observed among omnivores.

**Figure 2.**
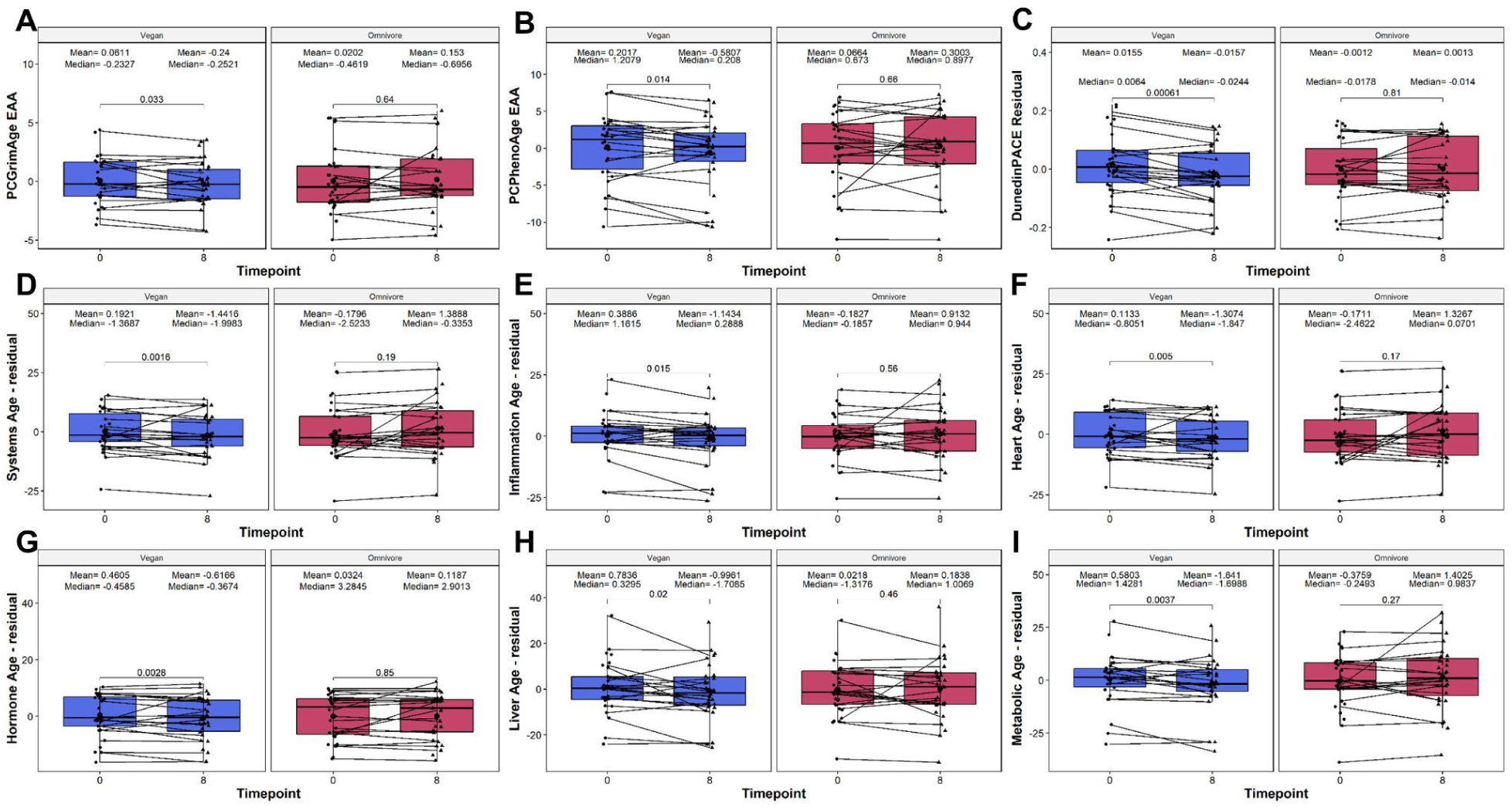
Boxplot showing the evolution of epigenetic age acceleration (EAA)/residuals among the significant epigenetic age clocks and systems-specific clocks based on diet type. Clocks assessed include the (A) PC GrimAge, (B) PC PhenoAge, (C) DunedinPACE, (D) Systems Age, (E) Inflammation Age, (F) Heart Age, (G) Liver Age, (H) Metabolic Age, and (I) Musculoskeletal Age. On the X-axis, the time points of measurements in weeks. On the Y-axis, the EAA/residual measure. EAA, or residual, is defined as the residual calculated between the raw value regressed upon chronological age, and adjusted by sex, technical principal components 1 and 2. On the top, the mean and median values of the EAA at each time point. The p-values of the paired Wilcoxon-rank sums test are also displayed in the plots. Lines that connect both boxplots represent the average of each patient’s tests.

### Analysis of cell cycle changes shows no significant changes based on diet

We next assessed whether diet type exhibited differences in overall mitotic rate as quantified by the mitotic clock output. Using the *epiTOC2* algorithm, we observed no significant changes in either the vegan or omnivore diet when assessing the total number of stem cell replication cycles estimated (*tnsc*) or the intrinsic stem cell cycle rate based on tissue (*irS*). This suggests that diet type did not have an impact on the overall mitotic clock values from the data.

### Vegan diets exhibit significant changes in relative Basophil levels

The immune system undergoes distinctive changes based on dietary choices, with vegan and omnivore diets influencing immune cell behavior in unique ways. Exploring this interplay provides valuable insights into the intricate relationship between diet and the body’s immune defenses. To investigate the impact of diet on the immune system, we next analyzed relative immune cell subset changes throughout the trial among 12 immune cell subsets as quantified by the EPIDISH frame: CD8T-naive, CD8T-memory, CD4T-naive, CD4T-memory, Basophils, B naive, B memory, T-regulatory, Monocytes, Neutrophils, Natural Killer, and Eosinophils. We observed significant changes in basophil levels in the vegan and omnivore diets. However, the basophil levels increases in the vegan group (delta mean = 0.0014, p = 0.04, **Figure 3**) and decreased in the omnivore group (delta mean = -0.0018, p = 0.048).

**Figure 3.**
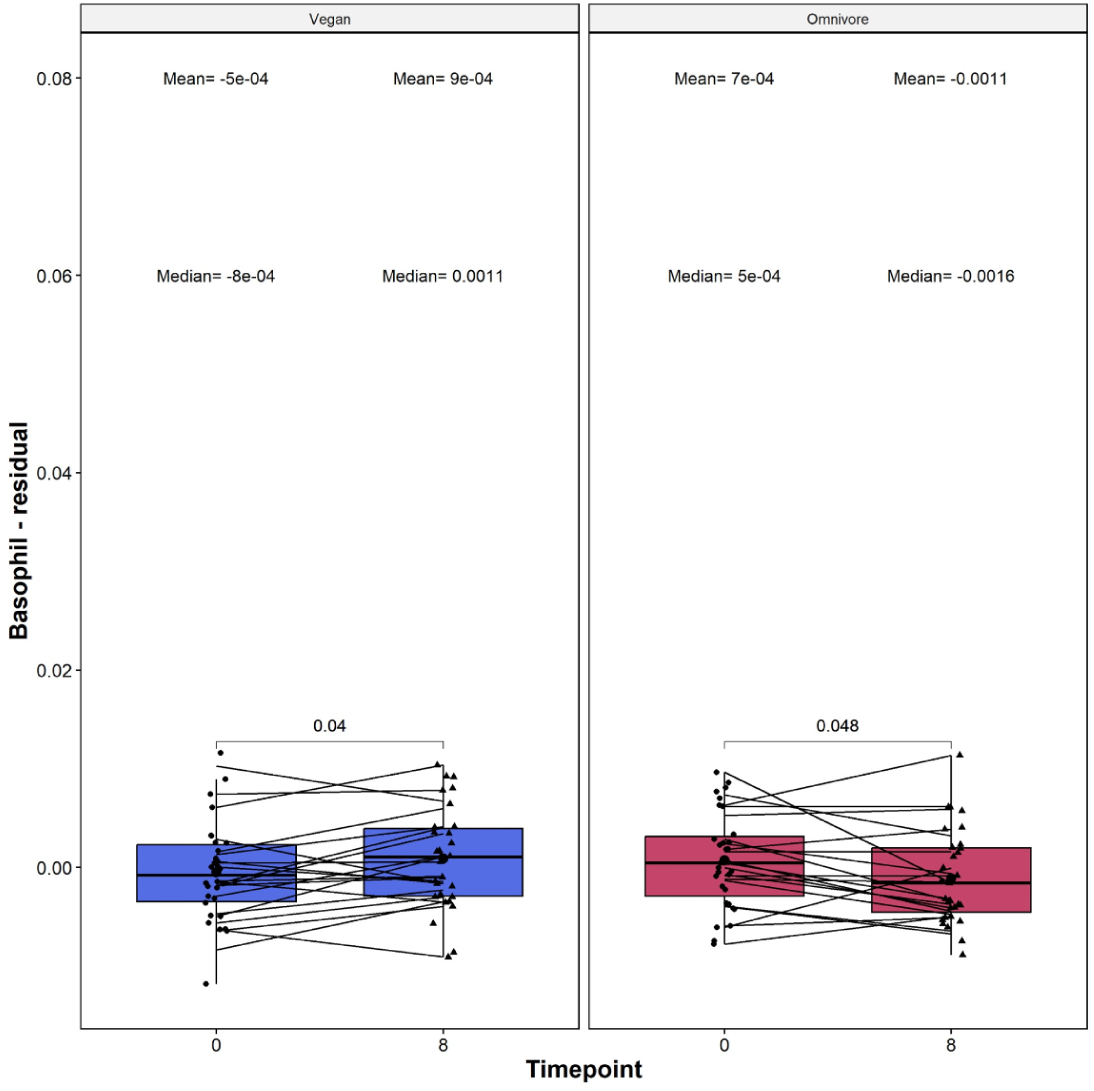
Boxplots showing the evolution of basophil cell subset percentages based on diet type. On the X-axis, the time points of measurements in weeks. On the Y-axis, the Basophil measure. The basophil measure is residualized, which is defined as the residual of the raw deconvolution value regressed upon chronological age, and adjusted by sex, technical principal components 1 and 2. On the top, the mean and median values of the residual at each time point. The p-values of the paired Wilcoxon-rank sums test are also displayed in the plots. Lines that connect both boxplots represent the average of each patient’s tests.

### Assessment of Type-2 Diabetes risk based on loci

We analyzed two DNA methylation loci, *ABCG1* (cg06500161) and *PHOSPHO1* (cg02650017), which are implicated in predicting T2D risk. Increased methylation in *ABCG1* correlates with a higher T2D risk, while heightened *PHOSPHO1* methylation is linked to a reduced risk. In our study, the vegan group displayed a noteworthy increase in cg06500161 methylation (delta beta value mean = 0.0105, p = 0.0093, see **Figure 4A**), indicating a potentially elevated T2D risk. Concurrently, an increase in cg02650017 methylation (delta beta value mean = 0.0079, p = 0.011, see **Figure 4B**) suggests a decreased T2D risk for the vegan cohort. This dichotomy in methylation changes for the two loci within the vegan group underscores a complex relationship between diet and T2D biomarkers, necessitating further investigation for a comprehensive understanding. None of these CpG sites were differentially methylated in the omnivore group.

**Figure 4.**
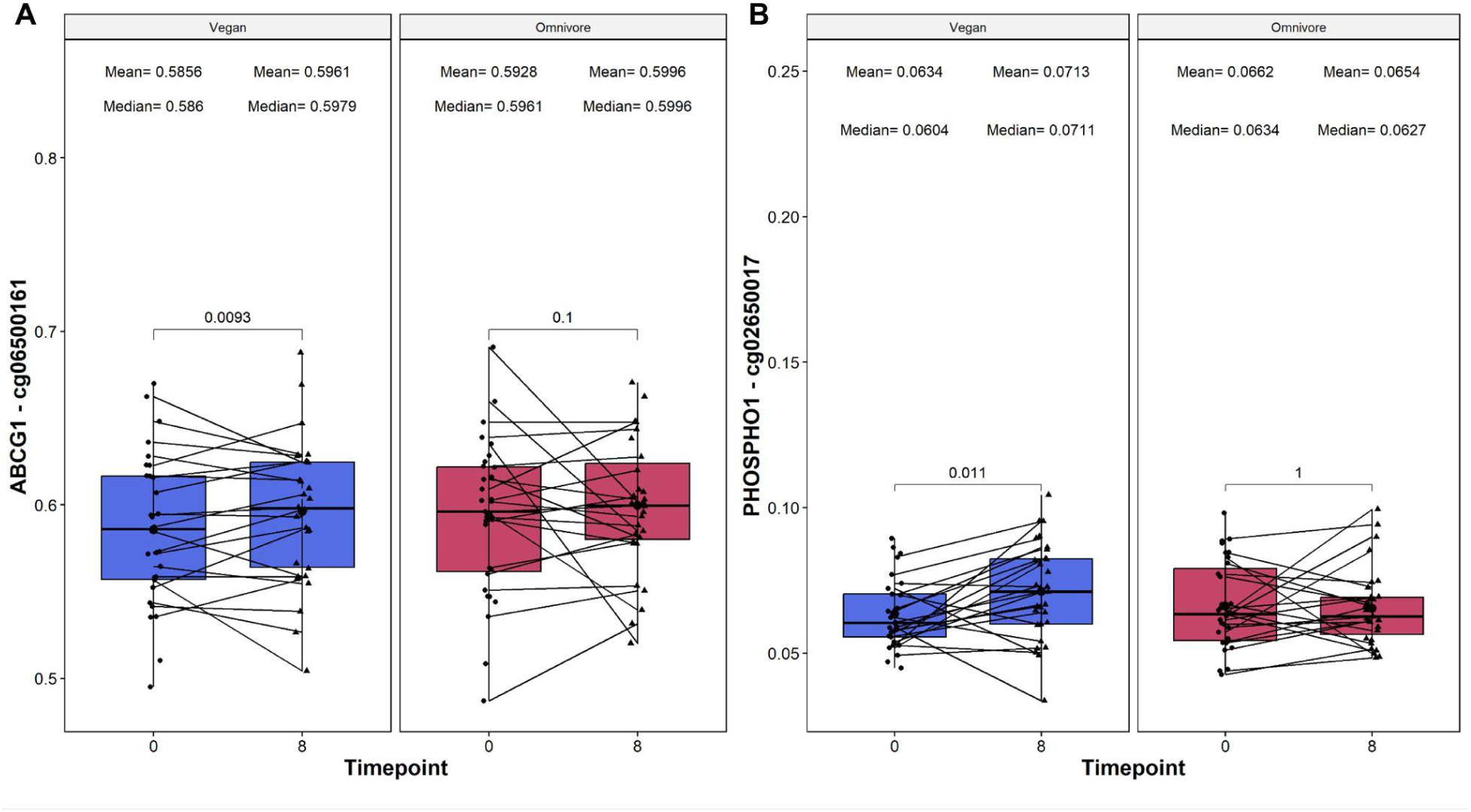
Boxplots showing the relative beta value change of two weight loss methylation sites on the ABCG1 gene (reported on the left) and PHOSPHO1 gene (reported on the right). On the X-axis, the time points of measurements in weeks, and the loci beta value which is reported on the Y-axis. On the top, the mean and median beta values of the loci at each time point. The p-values of the paired Wilcoxon-rank sums test are also displayed in the plots. Lines that are connecting both boxplots represent the average of each patient’s tests.

### Analysis of EpiScore markers

Recent efforts have expanded DNA methylation to predict proteins, complex behavioral, and physiological traits (McCartney et al. 2018; Gadd et al. 2022; Chen et al. 2023). To this end, we utilized these DNAm-based surrogate markers to assess relative changes in response to diet type (**Supplementary File S1**). In an initial analysis, we utilized 116 EpiScores that were previously described. In the Vegan group, we detected significant changes in seven EpiScores, namely CCL21, MMP1, ENPP7, Testican 2, ADAMTS, CD163, and MMP2, which were not evident in the omnivore group, underscoring diet-specific variations. Conversely, in the omnivore-specific analysis, six EpiScores—Ectodysplasin A, PAPP-A, VEGFA, HGF, Body Fat %, and TNFRSF17—exhibited exclusive significance. However, it must be noted that in both analyses, none of the EpiScore values met an adjusted significance threshold (adjusted p < 0.05); only the unadjusted. In summary, the methylation based surrogate markers of complex physiological and behavioral traits identified here suggest that while common markers are present, the majority of changes among the EpiScore are unique among diet types.

### Analysis of Epigenetic Biomarker Proxies (EBP)

We also assessed changes in EBPs, DNAm proxy scores of 396 metabolites, proteins, and clinical values (Chen et al. 2023). Of the 396, we identified a total of 76 and 89 EBPs which showed significant changes among the vegans and omnivores, respectively (**Supplementary File S2**). Independent analyses were performed between vegan and omnivore diets respectively to identify EBPs which showed: 1) unique changes among diet types, 2) consistent changes among diet types, and 3) opposing changes among diet types.

We identified 33 EBPs that showed uniquely significant changes (unadjusted p < 0.05) among the vegan cohort: *androsterone glucuronide, homovanillate (HVA), branched-chain, straight-chain, or cyclopropyl 10:1 fatty acid (2)*, Liver albumin, CCL18, PON1, dehydroepiandrosterone sulfate (DHEA-S), PON1, glutamine_degradant, leucine, 1,5-anhydroglucitol (1,5-AG), CRP, arabitol/xylitol, retinol (vitamin A), 3-hydroxyindolin-2-one sulfate, 2-methylcitrate/homocitrate, deoxycholic acid glucuronide, 7-hydroxyindole sulfate, alpha-CMBHC glucuronide, PCOC1, riboflavin (vitamin B2), 1-palmitoyl-GPC (16:0), PCOC1, GRN, S-carboxyethylcysteine, FETUA, CSPG2, dimethyl sulfone, carotene diol (2), guanidinosuccinate, 6-oxopiperidine-2-carboxylate*. Among these, 3 EBPs - *androsterone glucuronide, homovanillate (HVA), branched-chain, straight-chain, or cyclopropyl 10:1 fatty acid (2)\** - further passed an adjusted p-value threshold of 0.05, suggesting that the EBPs identified here represent potential biomarkers uniquely altered in response to a vegan diet at 8 weeks.

Among omnivores, we observed 46 EBPs which showed significant changes only among the omnivore diet cohort: *4-methoxyphenol sulfate, N-methylpipecolate, N-acetylcitrulline, sucrose, vanillactate, uridine, N-acetyltyrosine, 3-hydroxybutyroylglycine**, Liver_ALP, tryptophan, dihydroferulic acid sulfate, salicyluric glucuronide*, picolinate, 3,5-dichloro-2,6-dihydroxybenzoic acid, urea, galactonate, thyroxine, 2-acetamidophenol sulfate, cystathionine, sphinganine-1-phosphate, choline phosphate, picolinoylglycine, N,N,N-trimethyl-5-aminovalerate, 1-pentadecanoyl-GPC (15:0)*, TLL1, PCOC1, glycochenodeoxycholate 3-sulfate, trans-4-hydroxyproline, gentisate, catechol glucuronide, citramalate, ferulic acid 4-sulfate, PLMN, sedoheptulose, vanillic acid glycine, PCOC1, BMP1, linoleoylcarnitine (C18:2)*, 1-methylguanidine, isobutyrylcarnitine (C4), indolebutyrate, hypoxanthine, Smoking_PackYears, 3-hydroxyoctanoylcarnitine (1), eicosenoylcarnitine (C20:1)*, and BMP1*. Among these, 8 EBPs passed an adjusted p-value threshold, with *4-methoxyphenol sulfate, N-methylpipecolate, N-acetylcitrulline, sucrose, vanillactate, uridine* and *N-acetyltyrosine* exhibiting a significant increase among the omnivore group at Week 8, and a significant decrease in *uridine* and *3-hydroxybutyroylglycine*. These EBPs represent biomarkers uniquely associated with the omnivore diet but not vegan diet.

We also identified several EBPs which showed consistent changes among the diet types. Approximately 16 of the vegan EBPs showed significant increase in both vegan and omnivore diet types which included *CCL16, glucuronide of C12H22O4 (2)*, 2-methoxyhydroquinone sulfate (1), adenosine, lactosyl-N-palmitoyl-sphingosine (d18:1/16:0), 1-stearoyl-2-dihomo-linolenoyl-GPC (18:0/20:3n3 or 6)*, N-acetylalliin, N-carbamoylalanine, caffeine, carnitine, 1-palmitoyl-2-arachidonoyl-GPE (16:0/20:4)*, FETUA, 2,3-dihydroxy-2-methylbutyrate, LYSC, eicosenedioate (C20:1-DC)*, and 1-methyl-5-imidazoleacetate)*. Conversely, approximately 21 exhibited decreases among both diets, which included *10-undecenoate (11:1n1), 1,2-dipalmitoyl-GPC (16:0/16:0), 3-carboxy-4-methyl-5-propyl-2-furanpropanoate (CMPF), salicylate, succinylcarnitine (C4-DC), 1-margaroyl-2-arachidonoyl-GPC (17:0/20:4)*, 5-methyluridine (ribothymidine), Glucose, 2-aminoheptanoate, stearoyl-arachidonoyl-glycerol (18:0/20:4) [1]*, PCOC1, proline, ibuprofen, 11-ketoetiocholanolone glucuronide, homoarginine, Triglyceride, PCOC1, PCOC1, 1-stearoyl-2-adrenoyl-GPC (18:0/22:4)*, BMI, and 3-hydroxyphenylacetoylglutamine*). These EBPs represent metabolite, clinical, and protein targets which changed regardless of diet type, suggesting these as non-diet associated targets.

However, we observed 6 EBPs which showed opposing changes in EBP levels: *serine, 1-margaroyl-GPE(17:0)*, and 4-acetamidophenol,* showed significant increases among vegans, and significant decreases among omnivores, while *ergothioneine, indoleacetylglutamine,* and *creatinine* showed a significant decrease among vegans compared to the increase observed among omnivores. The significant, and opposing, changes between diets suggest that these represent diet based interactions significant in one diet but not the other.

Lastly, we also assessed the reproducibility of the EBPs to clinical outputs, compared the BMI-EBP changes relative to the BMI-clinical values that were collected within this study. We observed that both BMI measurements showed consistent significant decreases in both diet types. However, the magnitude of change was higher in the BMI-clinical values compared to the BMI-EBP values (**Figure 5**). Taken together, these findings exhibit the reproducibility of the BMI metrics among the EBPs relative to their clinical counterparts.

**Figure 5.**
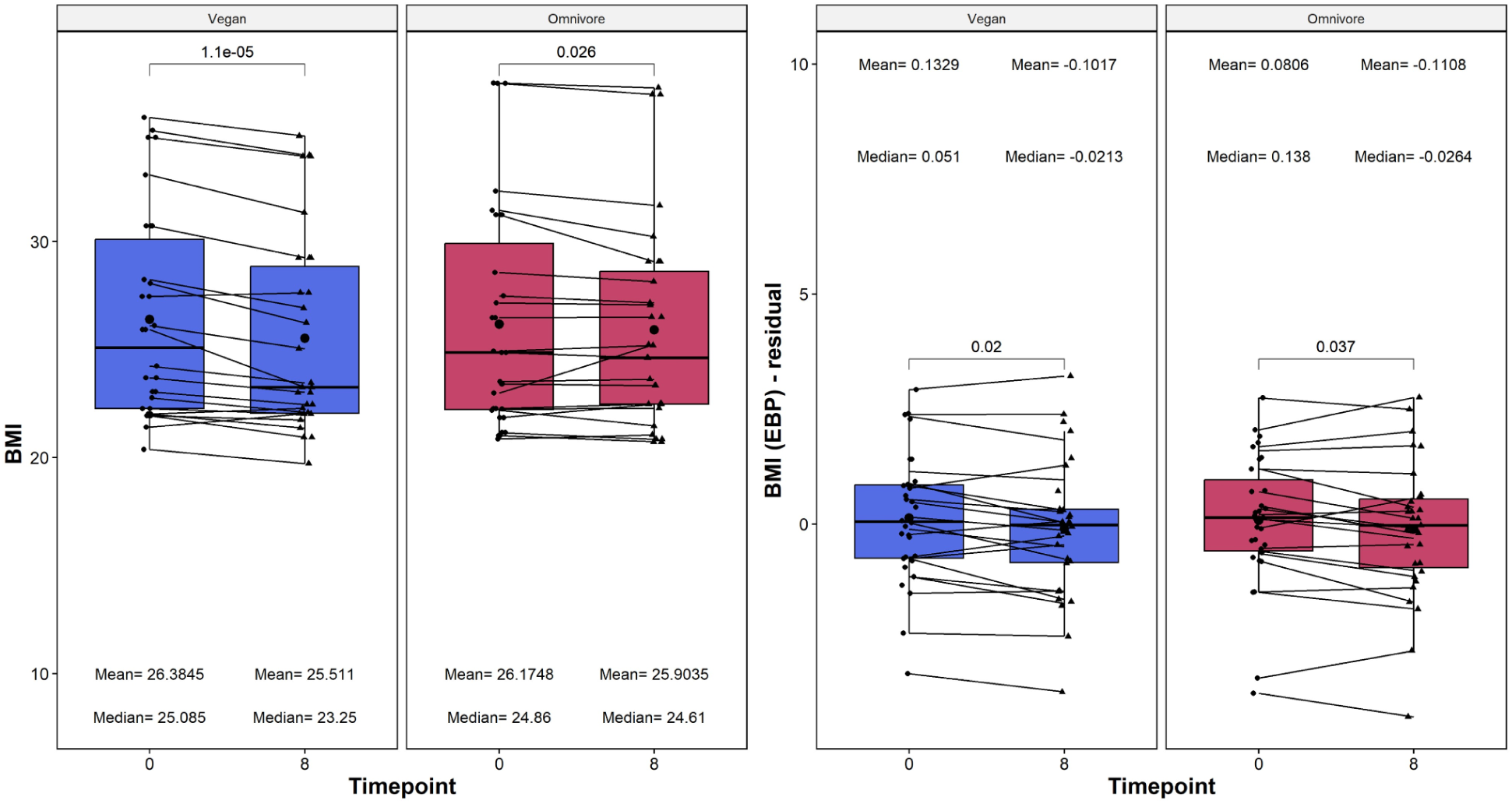
Boxplot showing the evolution of BMI values calculated from clinical measures (reported on the left) and epigenetic biomarker proxy (EBP) measures (reported on the right). On the X-axis, the time points of measurements in weeks. On the Y-axis, the BMI measure. The BMI-EBP measurements are reported as residuals, which are defined as the residual of the raw BMI value regressed upon chronological age, and adjusted by sex, technical principal components 1 and 2. No residual calculation was done for the clinical EBP. On the top, the mean and median values of the BMI at each time point. The p-values of the paired Wilcoxon-rank sums test are also displayed in the plots. Lines that connect both boxplots represent the average of each patient’s tests.

### Global EWAS analysis identifies epigenetic markers of vegan and omnivorous diets

Analysis of global DNA methylation patterns across the entire epigenome revealed differential methylation profiles between the experimental groups. Among the vegan cohort, we identified a total of 607 Differentially Methylated Loci (DMLs) across the genome (unadjusted p-value < 0.001) (**Figure 6A**). Among these, 322 CpG sites showed hypomethylation at 8 weeks, and 312 loci exhibited hypermethylation at Week 8 in response to the vegan diet. Among the omnivore cohort, a total of 494 DMLs were identified across the genome (**Figure 6B**), in which 309 CpGs exhibited significant hypermethylation and 185 CpGs exhibited hypomethylation at Week 8. The full list of CpGs is listed in **Supplementary File S3** for both analyses.

**Figure 6.**
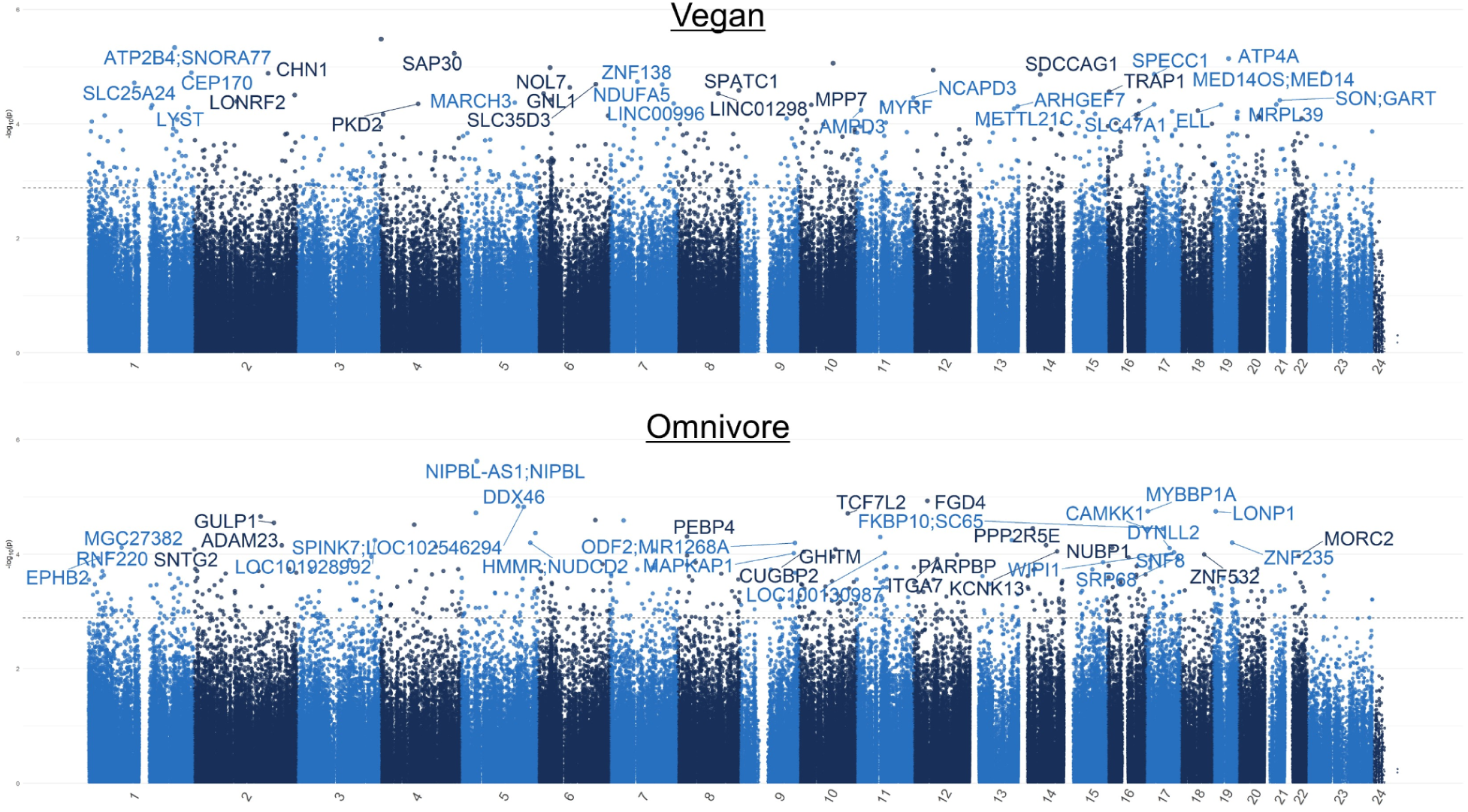
Manhattan plots for the vegan and the omnivore epigenome-wide association studies. The Manhattan plot illustrates genes associated with CpG sites identified in the (A) vegan and (B) omnivore comparison, with each dot representing a CpG site and its vertical position corresponding to the negative logarithm (base 10) of the unadjusted p-value for DNA methylation association (significance set at p = 0.001). The x-axis denotes genomic positions organized by chromosomes, with color-coded dots indicating specific chromosomes, and prominently peaked dots represent significantly associated CpG sites surpassing the genome-wide significance threshold.

To better understand the specific methylation patterns that differentiated vegan diet samples and omnivorous diets, a final analysis comparing the week 8 time points was conducted. We identified a total of 980 DMLs that were differentially methylated between the participants on an omnivore diet at week 8 and the participants on a vegan diet at week 8. Of the DMLs identified, 317 showed higher methylation in the vegan, and 663 DMLs exhibited higher methylation values in the omnivore group (**Figure 7A, Supplementary File S4**).

**Figure 7.**
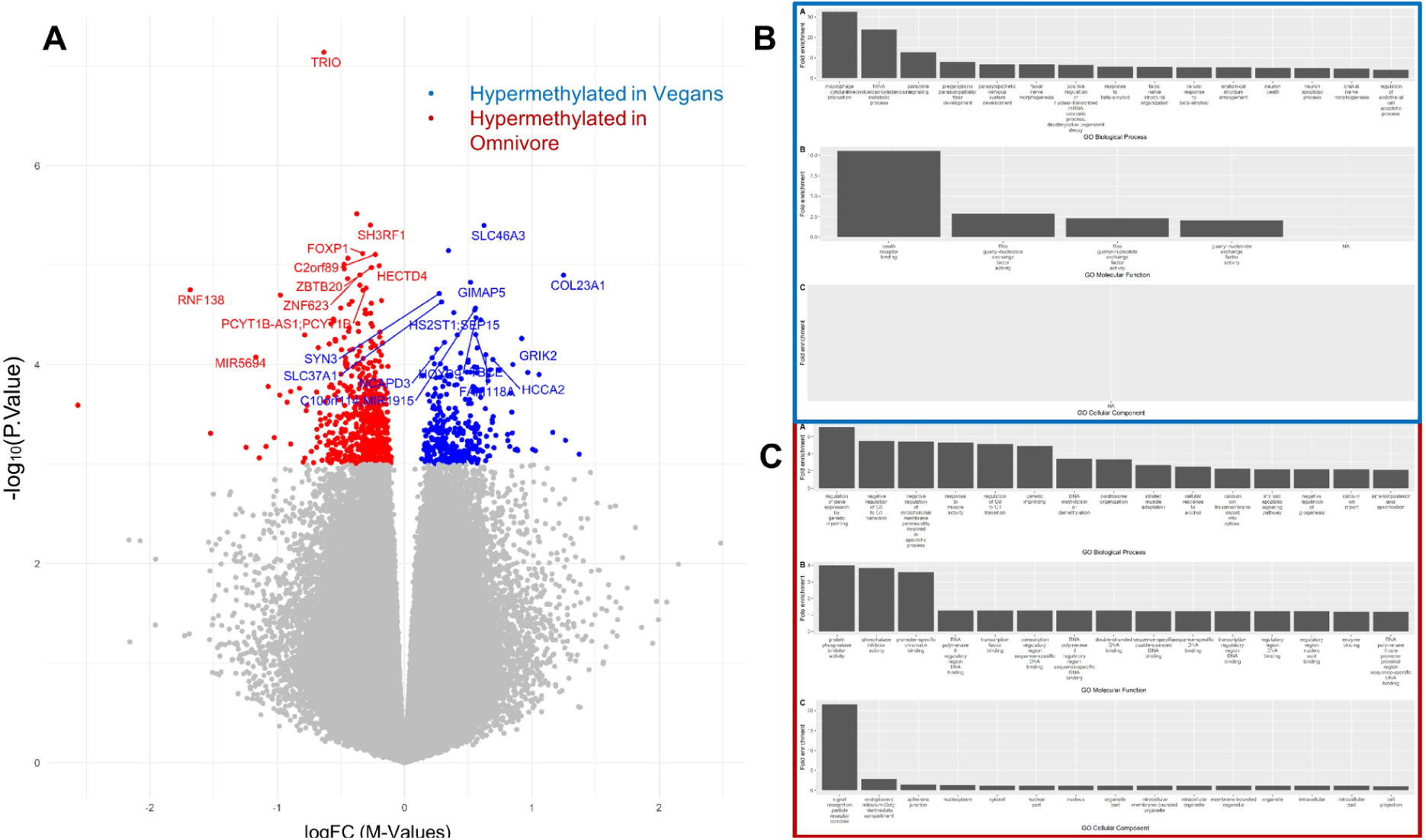
Epigenome-wide association study and enrichment analysis of the comparison between vegan and omnivore diet at the week 8 timepoint. (A) The Volcano plot illustrates DMLs identified in the Vegan vs. Omnivore comparison, with each dot representing a CpG site and its vertical position corresponding to the negative logarithm (base 10) of the unadjusted p-value for DNA methylation association. The x-axis denotes the relative log fold change (logFC) of the m-values between the vegan and the omnivore diets. Values greater than 0 represent CpGs with greater methylation among vegans (blue), compared to the negative values which represent greater methylation among omnivores (red). The biological associations for the hypermethylated CpGs among vegans are represented in (B), while hypermethylated CpGs among omnivores are represented in (C). Biological associations shown are gene ontology (GO) terms for Biological Processes (BP), Molecular Function (MF), and cellular components (CC).

To link the association of methylation to biological processes, enrichment analyses using the GREAT software were conducted on CpGs based on the direction of methylation. CpGs exhibiting significant hypermethylation in the vegan group, or hypomethylated among omnivores, were significantly enriched for gene-ontology biological process (GO-BP) terms such as *paracrine signaling, response to beta-amyloid, neuron apoptosis,* and several developmental GO-BP terms (**Figure 7B**). In addition, molecular function (GO-MF) terms such as *Ras guanyl-nucleotide exchange factor activity* were enriched for sites that exhibited significant hypermethylation among the vegans, and lower among the omnivores (**Supplemental File S5**). CpGs that were hypermethylated among omnivores, and hypomethylated among vegans, were enriched for GO-BP terms associated with cell cycle (negative regulation of the G0-G1 transition), genomic imprinting (*regulation of gene expression by genetic imprinting*), *cytosolic calcium ion transport,* and *cellular response to alcohol.* Cell cycle and transcriptional activity were further supported by the enrichment of GO-MF terms associated with RNA polymerase activity and transcriptional processes (*protein phosphatase inhibitor activity, RNA polymerase II regulatory region DNA binding,* and *promoter-specific chromatin binding*). Full results for biological associations to CpGs differentially methylated between diet types are listed in **Supplementary Table S6.** In summary, differential methylation patterns between vegan and omnivore groups reveal distinct associations with key biological processes and molecular functions, shedding light on the epigenetic mechanisms influencing diverse aspects of cellular activity in response to dietary choices.

## Discussion

In this study, we sought to elucidate the impact of a “healthy vegan” or a “healthy omnivorous diet” on epigenetic age, telomere length, immune cell subsets, and type 2 diabetes (T2D) risk-associated CpGs, building on current knowledge of nutrition on both diets. Our findings reveal distinct responses to vegan and omnivore diets, aligning with existing literature on the subject. Notably, the vegan cohort exhibited a significant decrease in epigenetic age acceleration, as demonstrated by reductions in multiple epigenetic aging clocks, all of which were trained upon clinical and phenotypic scores (PC GrimAge, PC PhenoAge, and DunedinPACE). The usage of systems-specific aging predictors further specified which organ systems showed age improvements, resolving five specific systems that showed aging improvements among the vegan cohort and not omnivores. These findings are consistent with previous research highlighting the potential anti-aging effects of plant-based diets, known for their rich antioxidant content and anti-inflammatory properties (Wang et al. 2023; Smith et al. 2020; Ornish et al. 2013). However, the significant impact of basophils in the vegan group contrasts with studies emphasizing the immunomodulatory benefits of plant-based diets, suggesting that further investigation into the nuanced interactions is warranted (Turner-McGrievy et al. 2015). These comprehensive findings underscore the complex interplay between diet, epigenetic regulation, immune function, and metabolic health, offering valuable insights for future research and personalized health interventions.

The measures investigated in our study offer a holistic perspective on biological aging without isolating system-specific aging processes, as highlighted by Ahadi in 2020 (Ahadi et al. 2020). However, the incorporation of the systems age clock in our research addresses this limitation by providing valuable, system-specific insights into aging changes (Sehgal et al. 2023). Notably, our findings reveal significant reductions in key physiological systems, including inflammation, heart, liver, metabolic, and hormonal systems. This nuanced approach aligns with previous research demonstrating that vegan and plant-based diets are associated with lower levels of inflammatory markers (Satija et al. 2016), lower risk of cardiovascular diseases (Orlich et al. 2013; H. Kim et al. 2019), reduced risk of non-alcoholic fatty liver disease (NAFLD) (Lv et al. 2023), improve glycemic control and other metabolic factors in individuals with type 2 diabetes (Barnard et al. 2009), and regulated hormonal level outputs in responses such as hot flashes (Kahleova et al. 2023). This approach allows for a more comprehensive understanding of the impact of the studied interventions on specific facets of aging, shedding light on potential areas of targeted intervention for promoting overall health and longevity. The identification of these system-specific changes contributes to a more nuanced and actionable comprehension of the aging process, underscoring the significance of our results in advancing our knowledge of interventions that may influence distinct physiological systems and enhance overall well-being.

While there is no gold standard measure of biological aging (Ferrucci et al. 2020), we analyzed several measures that represent the current DNAm predictors of biological aging. Nevertheless, these measures are acknowledged to be incomplete summaries of biological changes that occur with aging and to have technical limitations (Belsky et al. 2018; Bell et al. 2019). Treatment effects on aspects of biological aging not captured by the DNAm measures are not included in effect estimates; measurement error due to technical limitations of DNAm assays may bias effect estimates towards the null. Treatment effect estimates may therefore represent a lower bound of the true impact of vegan or omnivore dietary intervention on biological aging.

In our investigation of Epigenetic Biomarker Proxies (EBP), the notable consistency in significant decreases observed in both BMI-EBP and BMI-clinical values across diet types highlights the reproducibility of BMI metrics within the epigenetic context. Despite a slightly higher magnitude of change in BMI-clinical values, the parallel trends emphasize the reliability of BMI-EBPs as reflective markers of body mass index alterations. Remarkably, six Epigenetic Biomarker Proxies (EBPs) exhibited divergent alterations between the vegan and omnivore diets, shedding light on diet-specific impacts on the epigenome. Ergothioneine, indoleacetylglutamine, and creatinine demonstrated a noteworthy decrease in the vegan group but an increase in the omnivore group. Ergothioneine, a potent antioxidant guarding cells against oxidative stress, potentially decreased in the vegan diet due to reduced intake from sources like mushrooms and certain grains. Indoleacetylglutamine, derived from tryptophan, showcased elevated levels in the omnivore diet and a decline in the vegan diet, possibly mirroring the distinct abundance of protein-rich foods in each diet. The analogous patterns in creatinine, a marker of muscle metabolism, might also be linked to variations in protein intake and muscle turnover between the two diets. Conversely, serine, 1-margaroyl-GPE(17:0), and 4-acetamidophenol saw a significant rise in the vegan group but a decrease in the omnivore group. Serine, a non-essential amino acid abundant in plant sources, such as soybeans and nuts, likely increased on the vegan diet due to elevated consumption. The opposite trends in 1-margaroyl-GPE(17:0), a relatively novel metabolite predicted to function as a glycerophospholipid involved in cellular membranes and signaling pathways, suggest diet-induced variations in membrane composition and function. 4-acetamidophenol, a derivative of paracetamol widely used in analgesic and antipyretic medications, may reflect increased usage in the vegan compared to the omnivore group. Further studies are needed to identify the health implications of these changes and whether specific dietary components are responsible for them. The EpiScore analysis provided valuable insights into the potential of DNA methylation markers for predicting complex physiological and behavioral traits influenced by diet. Seven EpiScores showed significant changes exclusively in the vegan group, while six exhibited exclusive significance in the omnivore group. Although none met an adjusted significance threshold, the unadjusted values highlight the potential of DNA methylation-based surrogate markers in delineating diet-related impacts on complex traits. This underscores the necessity for further exploration to refine and validate these markers for their predictive utility.

Several metabolites EBPs exhibited noteworthy changes, providing insights into differences and commonalities of diet response between the two groups. Among the top markers showing significant alterations in the vegan group, C-reactive protein (CRP), Deoxycholic Acid Glucuronide, and Spermidine stood out. A decrease in predicted CRP levels suggests a potential reduction in systemic inflammation. Spermidine, a polyamine associated with various health benefits, demonstrated an increase, potentially indicating an increased intake of vegetables like soy, legumes, and mushrooms. Deoxycholic Acid Glucuronide, a bile acid metabolite, displayed a decrease, suggesting an expected potential reduction in bile acid metabolism in response to a reduced intake of animal fat. Additionally, the vegan group demonstrated significant changes in other markers, such as N-acetyl-cadaverine and carnitine. Whereas the elevated levels of N-acetyl-cadaverine decreased as expected, given that this marker is associated with amino acid fermentation in the gut, the increase in carnitine levels contradicts the anticipated decrease in response to a vegan diet, since carnitine is mainly derived from meat and dairy products (Lombard et al. 1989).

Several metabolites exhibited significant decreases in both diet groups, pointing to shared metabolic responses across diverse dietary patterns. Both salicylate, a component found in various plant foods, and its metabolite salicyluric glucuronide, demonstrated a reduction in both groups potentially reflecting a decrease in salicylate rich food such as legumes (e.g., lentils, beans), vegetables (e.g., cauliflowers, pickled vegetables), and fruits (e.g., strawberries, plums, watermelons). Reductions in quinate, a compound derived from the metabolism of coffee polyphenols (Guertin et al. 2015; Playdon et al. 2016) and 10-undecenoate (11:1n1), a fatty acid related to butter intake (Pallister et al. 2016; Guertin et al. 2014) suggest potential reduction in coffee and butter intake, respectively. Interestingly, both groups exhibited a decrease in predicted body mass index (BMI), which is consistent with the decrease in BMI in both groups.

In the omnivore group, we observed several intriguing shifts in key metabolic markers. The increase in tryptophan and serotonin, a neurotransmitter synthesized from tryptophan, suggests potential impacts on mood regulation and other serotonin-mediated functions in response to increased intake of tryptophan-rich animal protein in the omnivore diet. Choline phosphate, a vital component in cell membrane structure, exhibited an increase, hinting at increased dietary intake from meat, fish, and eggs. Indolebutyrate, a microbial metabolite, displayed an increase, suggesting potential shifts in gut microbial metabolism influenced by the diverse dietary components. Adenosine, a nucleoside that promotes sleep and reduces anxiety, exhibited an increase, indicating potential changes in endogenous metabolism on an omnivore diet (Masino et al. 2009). These findings underscore the nuanced interplay of neurotransmitter synthesis, lipid metabolism, microbial activity, and purine metabolism associated with omnivorous dietary patterns.

Previous studies have suggested vegan diets associated with lower T2D risk (Satija et al. 2016; Tonstad et al. 2013). Interestingly, our investigation into T2D risk-associated methylation loci revealed that the vegan diet led to increased methylation in *ABCG1* and *PHOSPHO1*, which provided relatively conflicting results; increase in *ABCG1* indicates a reduced T2D risk, which is contradicted with the increase in *PHOSPHO1*, which indicates increased T2D risk. These results call for the need to develop disease-specific epigenetic predictors for T2D risk which go beyond single loci risk predictors, to potential multi-loci risk predictors exhibit significant association to disease risk.

Finally, the exploration of global DNA methylation patterns across the entire epigenome revealed notable disparities between the vegan and omnivore cohorts, identifying 607 and 494 differentially methylated loci (DMLs) across the genome, respectively. This comprehensive epigenome-wide analysis aligns with a growing body of literature examining the epigenetic effects of different dietary patterns (Do et al. 2021; Hellbach, Sinke, et al. 2023; Hellbach, Freuer, et al. 2023). When we analyzed each diet group independently, we observed 322 hypomethylated probes in the vegan diet and 185 in the omnivore diet. These CpG sites represent the epigenetic targets that changed during the trial, but independent of diet. However, to compare the evolution of each of the twin pairs, we compared the week 8 time point for those individuals in the vegan diet and those in the omnivore diet. This analysis unraveled 980 DMLs, with 317 demonstrating higher methylation in the vegan group and 663 in the omnivore group. Using the significant CpG sites from the twin-pair comparison at week 8, we performed an enrichment analysis to elucidate the biological relevance of these methylation patterns. Hypermethylated sites in vegans revealed enrichment of paracrine signaling, response to beta-amyloid, neuron apoptosis, and developmental processes. These findings imply that a vegan diet may influence pathways associated with cellular communication, neuroprotective mechanisms, and development. In contrast, hypermethylation in omnivores was linked to cell cycle regulation, genomic imprinting, cytosolic calcium ion transport, and cellular response to alcohol. This suggests that an omnivorous diet may impact pathways related to cell division, genetic regulation, cellular signaling, and responses to environmental stimuli. These insights contribute to a deeper understanding of how diet can impact the epigenome and, consequently, influence various aspects of cellular activity and health outcomes. Future investigations linking the epigenetic sites identified here in the context of gene expression may identify gene regulatory networks altered due to diet, further providing a molecular perspective in nutrition and diet.

It is crucial to acknowledge that the observed epigenetic age and biomarker differences between the vegan and omnivore groups may be predominantly attributed to the variations in weight loss rather than solely reflecting the distinct dietary compositions. Throughout the “Food Delivery” phase, the vegan group consumed ∼ 200 calories less per day than their omnivorous counterparts, resulting in an average weight loss of 2 kilograms greater than the omnivore group by the end of the 8-week intervention. Extensive population studies and Mendelian randomization analyses have underscored the impact of BMI changes on inducing epigenetic alterations linked to metabolic health (Wahl et al. 2017; Mendelson et al. 2017). However, it should be noted that while we saw significant decreases in both clinical-BMI and EBP-BMI values, only the vegan cohort exhibited significant reductions in epigenetic age. This calls for a nuanced interpretation of our findings and emphasizes the need for future investigations to disentangle the complex interrelationships between dietary factors, weight dynamics, and epigenetic modifications.

Our study provides valuable insights into the short-term effects of weight loss on two different diets on epigenetic markers. However, it is important to acknowledge that the long-term impact of a vegan diet on epigenetic processes may carry adverse effects in the absence of sufficient intake of crucial vitamins and nutrients essential for supporting these intricate molecular reactions. In particular, all vegans and a substantial portion of vegetarians, if not supplemented, are at risk of developing vitamin B12 deficiency, resulting in elevated levels of homocysteine—an established marker of dysfunctional methylation associated with increased cardiovascular risk, including coronary artery disease (CAD) and heightened stroke susceptibility (Pawlak 2015; Tong et al. 2019; Kwok et al. 2005). Vitamin B12 deficiency has been implicated in disease-related epigenetic alterations in both animals and humans (Adaikalakoteswari et al. 2015; Waterland and Jirtle 2003; Mahajan et al. 2019; Ge, Zadeh, and Mohamadzadeh 2022). In our cohort, the vegan group exhibited a lower intake of vitamin B12, although serum vitamin B12 levels did not demonstrate statistical differences compared to omnivores at the 8-week mark, likely due to preserved stores (Landry et al. 2023). It is crucial to emphasize that long-term adherence to vegan diets typically necessitates vitamin B12 supplementation to mitigate the risk of deficiency and its consequential impact on epigenetic processes. This highlights the imperative role of nutritional considerations in optimizing the health outcomes associated with plant-based dietary choices. Within the context of these limitations, our findings have implications for future geroscience research. Aging biology research has identified multiple therapies with the potential to improve healthy lifespan in humans. A barrier to advancing the translation of these therapies through human trials is that intervention studies run for months or years, but human aging takes decades to cause disease (Sierra et al. 2021; Moffitt et al. 2017; Justice et al. 2016).

## Conclusions

In this epigenetic analysis of a randomized clinical trial, we observed significant changes among healthy identical twins which indicate potential advantageous aging benefits for the healthy vegan diet vs. the omnivorous diet. We further show the value of utilizing epigenetic testing to gain greater molecular aging insight in nutrition. Utilizing epigenetic tests in investigating diet types and nutrition is crucial as it offers a dynamic and personalized assessment of cellular aging, enabling targeted interventions to optimize dietary approaches and promote overall well-being.

## Methods

### Ethical Approval and Study Design

Procedures adhered to the ethical standards of the Helsinki Declaration, approved by the Stanford University Human Subjects Committee (IRB protocol 63955, approved March 9, 2022). Written informed consent was obtained from all participants. The study, a single-site, parallel-group dietary intervention trial, randomized generally healthy adult twins to either a healthy vegan or omnivorous diet for 8 weeks. Enrollment commenced in March 2022, concluding in May 2022, with the final follow-up in July 2022. The trial employed the CONSORT reporting guideline for randomized clinical trials, focusing on the primary outcome: the 8-week change in DNA methylation profiles from baseline. Secondary outcomes encompassed triglycerides, HDL-C, glucose, insulin, TMAO, vitamin B12, and body weight, serving as controls for relevant methylation risk scores. Diet quality, adherence, and study design are illustrated in **Figure 1**.

### Participant Recruitment and Eligibility

The goal was to recruit 22 pairs of identical twins, primarily from the Stanford Twin Registry and other twin registries, including Netflix’s pre-recruited participants interested in a documentary on vegan diets. Inclusion criteria involved participants aged ≥18, part of a willing twin pair, with BMI <40, and LDL-C <190 mg/dL. Exclusions included uncontrolled hypertension, metabolic disease, diabetes, cancer, heart/renal/liver disease, pregnancy, lactation, and medication use affecting body weight or energy. Eligibility was determined via online screening, followed by an orientation meeting and in-person clinic visit. Randomization occurred only after completing baseline visits, dietary recalls, and questionnaires for both twins. One twin pair ( which started the study, did not abide by the above requirements and thus was removed from the study. Ultimately blood samples from 21 numbers of twin pairs (N = 42) were considered for downstream analyses.

### Dietary Intervention

The study comprised two 4-week phases: delivered meals and self-provided meals. Trifecta Nutrition supplied meals for the first 4 weeks, tailored to omnivorous and vegan diets. Health educators facilitated nutrition classes via Zoom, emphasizing principles like choosing minimally processed foods and building balanced plates. The omnivorous group received animal product targets (e.g., 6-8 ounces of meat, 1 egg, and 1.5 servings of dairy), while the vegan group avoided all animal products. Dietary intake was assessed through unannounced 24-hour recalls and participant logs on the Cronometer app, capturing food intake details at baseline, week 4, and week 8. Dietary data quality was ensured through trained dietitian interviews and app records, used to evaluate diet quality and adherence.

#### DNA Methylation Assessment

Whole blood was collected at baseline and at week 8 for DNA methylation preparation and analysis. Majority of twin pairs (20 twin pairs, N = 40) were collected as biological replicates per time point and individual using Dried Blood Spot cards; one twin pair (N=2 patients) which had triplicate collections in which two were collected by dried blood spot and one using the tasso. Blood collected by the clinics was sent to TruDiagnostic labs in Lexington, KY, for DNA extraction and methylation processing. Using the EZ DNA Methylation kit (Zymo Research), 500 ng of DNA was bisulfite-converted following the manufacturer’s instructions. Bisulfite-converted DNA samples were randomly assigned to wells on the Infinium HumanMethylationEPIC BeadChip, and the subsequent steps included amplification, hybridization, staining, washing, and imaging with the Illumina iScan SQ instrument to acquire raw image intensities. Longitudinal DNA samples for each participant were assessed on the same array to mitigate batch effects. Raw image intensities were saved as IDATs for further processing.

#### DNAm Data Processing

Raw IDATs underwent processing using the *minfi* pipeline (Aryee et al. 2014). Samples of low quality were identified with ENMix based on variance of internal controls, flagging samples showing more than 3 standard deviations away from the mean control probe value (Xu, Niu, and Taylor 2021). However, no outlier samples were identified, and thus, all samples were considered for analysis. DNAm normalization involves Gaussian mixed quantile normalization (GMQN) to correct between batch collections and BMIQ normalization to normalize intra-sample variance within chips (Xiong et al. 2022). Probe-level analysis masked probe sets without at least one intensity fluorescence above the background. Missing beta values were imputed using K nearest neighbor (KNN) imputation.

#### Deriving Estimates of Epigenetic Clocks and Methylation-Based Metrics

Epigenetic clocks were calculated from cleaned beta values, focusing on clocks like Horvath multi-tissue (Horvath 2013), Horvath skin and blood (Horvath et al. 2018), Hannum (Hannum et al. 2013), PhenoAge (Levine et al. 2018), GrimAge v1 and v2 (Lu, Quach, et al. 2019; Lu et al. 2022) and DNAmTL (Lu, Seeboth, et al. 2019). To ensure that values were highly reproducible, the principal component versions of these clocks were utilized as described by Higgins-Chen et al. (Higgins-Chen et al. 2022). Individual systems clocks were calculated using the framework presented by Sehgal et al. (Sehgal et al. 2023). Clocks were calculated using a custom R script available on Github. DunedinPACE was calculated using a custom script available from Github (https://github.com/danbelsky/DunedinPACE, (Belsky et al. 2022)). Additional non-epigenetic age metrics included relative percentages of 12 immune cell subsets imputed using EpiDISH (Luo et al. 2023), 116 methylation-based predictions of biochemical and lifestyle risk factors using MethylDetectR (Hillary and Marioni 2020), and 396 epigenetic biomarker proxies (Chen et al. 2023). All epigenetic metrics such as clocks, telomere length, immune deconvolution, EpiScore, and EBPs, were residualized prior to statistical analysis by using the *lmer()* R package as such:

> *Residualized epigenetic metric = resid(lmer(Epigenetic predictor ∼ Chronological Age + Sex + PC1 + PC2)*

Statistical analyses were performed using paired Wilcoxon-rank sum tests faceted by diet type, with significance set at unadjusted p < 0.05.

#### Differentially Methylated Analysis

Differential methylation analysis was conducted using processed beta values logit-transformed to M-values with the *BetaValueToMValue* function from the *sesame* R package. *Limma* package was applied across three comparisons, identifying differentially methylated loci (DMLs) within each diet between baseline and week 8 and between omnivore and vegan diets at week 8. Multivariate linear models were controlled for fixed effects such as chronological age, BMI, sex, beadchip, 5 immune cell percentages (Basophils, CD8T naive, Eosinophils, NK, and Neu), the first three principal components of technical probes, and random effects such as twin pair designation in the vegan vs. omnivore comparison at week 8 and patient ID in the longitudinal comparisons. Similarly, for the longitudinal analysis of the independent vegan and omnivore comparisons, the same fixed effects and PC components were used, however the individual ID was used in the longitudinal comparison. The inflation or deflation of P-values across the methylome was assessed with Q-Q plots and lambda values (Guintivano et al., n.d.). We evaluated multiple models including different fixed effects and the one described was the model with a lambda closest to 1. DMLs were identified with a significance level of unadjusted p < 0.001. Functional annotation of DMLs was performed using the GREAT pipeline to identify significant gene ontology terms, as implemented in the *rGREAT* R package (Gu and Hübschmann 2023).

## Data Availability

All data produced in the presesnt study are available upon reasonable request to the corresponding authors.

## Author Contributions

VBD, LA, and CDG had full access to the data and verified the data integrity and accuracy of the analysis.

Writing of manuscript – VBD, LA, NCG, AL, LT, RS

Conceived and designed the study, and provided funding - CDG, JLS

Sample Selection, Patient Recruitment, and sample analysis– TH, JLR

Sample processing and Data generation - TLM

Data processing, normalization, statistical analysis, EWAS analysis - VBD Results interpretation - VBD, LA

Figure generation – VBD, NCG, AL, LT

Edited manuscript - all authors

## Conflict of Interest Disclosures

Dr. Dwaraka, Dr. Carreras-Gallo, Aaron Lin, Logan Turner, Dr. Mendez, Hannah Went, and Ryan Smith are all employees of TruDiagnostic Inc. Dr Gardner reported receiving funding from Beyond Meat outside the submitted work. Dr J. L. Sonnenburg is a Chan Zuckerberg Biohub investigator. No other disclosures were reported.

## Role of the Funder/Sponsor

The funders had no role in the design and conduct of the study; collection, management, analysis, and interpretation of the data; preparation, review, or approval of the manuscript; and decision to submit the manuscript for publication.

## Transparency Declaration

The lead authors, Varun B. Dwaraka and Lucia Aronica, (the manuscript’s guarantor) affirm that the manuscript is an honest, accurate, and transparent account of the study being reported; that no important aspects of the study have been omitted; and that any discrepancies from the study as planned (and, if relevant, registered) have been explained.

## Data Sharing Statement

The data that support the findings of this study are available from the corresponding authors upon reasonable request.

## Trial Registration

ClinicalTrials.gov Identifier: NCT05297825

## Supplemental Material

**<u>Supplemental File S1.</u> EpiScore analysis between baseline and 8-week test in the Stanford TWINs trial.** The first column reports the EpiScore that was assessed, followed by the unadjusted p-value, and the direction of difference of the residual values between Week 8 from Week 0 for the Vegan (columns 2 and 3), and omnivore samples (columns 4 and 5). The final two columns show adjusted p-values for the vegan (column 6) and omnivore analyses (column 7). The statistical test run here was the Wilcoxon-rank sum test. The direction of change is represented as a + (representing a higher value at Week 8 relative to Week 0) or a - (representing a lower value at Week 8 relative to Week 0). Abbreviations: NS = not significant.

**<u>Supplemental File S2.</u> Epigenetic Biomarker Proxy (EBP) analysis between baseline and 8-week test in the Stanford TWINs trial.** The first column reports the EBP that was assessed, followed by the unadjusted p-value, and the direction of difference of the residual values between Week 8 from Week 0 for the Vegan (columns 2 and 3), and omnivore samples (columns 4 and 5). The final two columns show adjusted p-values for the vegan (column 6) and omnivore analyses (column 7). The statistical test run here was the Wilcoxon-rank sum test. The direction of change is represented as a + (representing a higher value at Week 8 relative to Week 0) or a - (representing a lower value at Week 8 relative to Week 0). Abbreviations: NS = not significant.

**<u>Supplemental File S3</u>. Excel file contains the significant results for the differential methylation analysis results from the EWAS time point analysis of Week 8 vs Week 0 of the Vegan diet.** Column headers of each sheet are listed as follows: Column A represents the CpGs identified; Column B shows the log fold change of the m-value between Week 0 vs. Week 8 for each timepoint comparison, in which positive values are higher methylation at week 8 relative to week 0; Column C shows the average M-value for the CpG; Column D reports the t-statistic; Column E reports the unadjusted p-value; Column F reports the false-discovery rate (FDR) corrected p-value; Column G reports the B value outputted from limma; and Column H reports the gene ID overlapping the specific CpG loci.

**<u>Supplemental File S4.</u> Excel file contains the significant results for the differential methylation analysis results from the EWAS time-point analysis of Week 8 vs Week 0 of the Omnivore diet.** Column headers of each sheet are listed as follows: Column A represents the CpGs identified; Column B shows the log fold change of the m-value between Week 0 vs. Week 8 for each timepoint comparison, in which positive values are higher methylation at week 8 relative to week 0; Column C shows the average M-value for the CpG; Column D reports the t-statistic; Column E reports the unadjusted p-value; Column F reports the false-discovery rate (FDR) corrected p-value; Column G reports the B value outputted from limma; and Column H reports the gene ID overlapping the specific CpG loci.

**<u>Supplemental File S5.</u> Excel file contains the significant results for the differential methylation analysis results from the EWAS time-point analysis of the Week 8 Vegan compared to the Week 8 of the Omnivore diet.** Column headers of each sheet are listed as follows: Column A represents the CpGs identified; Column B shows the log fold change of the m-value between the Vegan vs. Omnivore at Week 8, in which positive values are higher methylation in the vegans relative to omnivores; Column C shows the average M-value for the CpG; Column D reports the t-statistic; Column E reports the unadjusted p-value; Column F reports the false-discovery rate (FDR) corrected p-value; Column G reports the B value outputted from limma; and Column H reports the gene ID overlapping the specific CpG loci.

**<u>Supplemental File S6</u>. GREAT results for DMLs hypermethylated in Vegan samples at 8 weeks, compared to Omnivore samples.** Columns presented are as follows: Column A represents the GO ID of the term identified, Column B exhibits the name of the GO ID, Column C exhibits the

**<u>Supplemental File S7</u>. GREAT results for DMLs hypermethylated in Omnivore samples at 8 weeks, compared to Vegan samples.**

## Notes

### Clinical Trial

NCT05297825

### Author Declarations

This study followed the ethical standards of the Declaration of Helsinki and was approved by the Stanford University Human Subjects Committee on March 9, 2022.

